# Clinician Suspicion for Lyme Disease and Clinical Decision-Making in Children with Monoarthritis

**DOI:** 10.64898/2026.05.06.26352605

**Authors:** Alexandra T. Geanacopoulos, Rebecca S. Green, Laura L. Chapman, Desiree N. Neville, Meagan M. Ladell, Amy D. Thompson, Anupam B. Kharbanda, Lise E. Nigrovic, Pedi Lyme Net

## Abstract

In this large multi-center cohort of children evaluated for Lyme disease in a Lyme-endemic emergency department, we assessed the diagnostic accuracy of clinician suspicion and subsequent clinical decision-making for children presenting with monoarthritis. Among 1,582 children with monoarthritis evaluated for Lyme disease, 623 (39%) had Lyme arthritis and 32 (2%) had septic arthritis. Overall, 313 (20%) had an invasive joint procedure (arthrocentesis or arthroscopy), 194 (12%) received parenteral antibiotics, and 376 (24%) were hospitalized. Clinician suspicion had moderate discriminative ability for Lyme disease (area under the receiver operating characteristics curve: 0.75, 95% confidence interval: 0.72-0.77). Children with higher clinician suspicion were less likely to receive parenteral antibiotics or to be hospitalized, although invasive procedure rates were similar. Our findings highlight the challenge of clinically distinguishing Lyme from septic arthritis. Better diagnostic tools are needed to improve timely diagnosis and minimize invasive testing among children with monoarthritis in Lyme-endemic regions.

## Introduction

Children with Lyme and septic arthritis can present similarly.^1^ While children with Lyme arthritis require oral antibiotics, those with septic arthritis need prompt surgical intervention and intravenous antibiotics to minimize damage to the joint space.^2^ Because current Lyme serologic testing takes days to result, initial decision-making often relies on clinician suspicion and imperfect blood biomarkers.^3, 4^ We evaluated the diagnostic accuracy of clinician suspicion for Lyme disease and subsequent clinical decision-making for children with monoarthritis from a high-incidence Lyme disease area.

## Methods

### Study design

We performed a secondary analysis of an institutional review board-approved prospective cohort of children (1 to 21 years) evaluated for Lyme disease in one of eight Pedi Lyme Net emergency departments (EDs, 2015-2024).^5^

### Study patients

For this sub-study, we selected children presenting with monoarthritis, defined by clinician-reported single joint swelling or hip pain with or without swelling, as hip swelling can be differentially assessed.^4^ We collected clinical and exam factors at the time of enrollment and performed phone and medical record review one month after enrollment.

### Definitions of Lyme and septic arthritis

We defined a case of Lyme arthritis with a positive two-tier serology test result obtained within 30 days of enrollment and septic arthritis with growth of pathogenic bacteria in synovial fluid culture or in blood culture paired with synovial fluid pleocytosis (≥50,000 white blood cells/high-powered field).^6^ All other children had inflammatory arthritis.

### Exposure and Outcomes

The primary exposure was clinician suspicion of Lyme disease (1 low to 10 high) recorded by the treating supervising clinician after completing initial history and physical examination, but before reviewing laboratory test results. Our outcomes were Lyme and septic arthritis diagnoses, completion of an invasive joint procedure (arthrocentesis or arthroscopy), initial receipt of parenteral antibiotics, and hospitalization.

### Analysis

We generated a receiver operating characteristics (ROC) curve to evaluate the diagnostic accuracy of clinician suspicion for Lyme arthritis using the area under the ROC curve (AUC). Bootstrap resampling (2,000 replicates) was used to generate 95% confidence intervals (CIs) across the ROC curve. We calculated the proportion and 95% Wilson CI of children diagnosed with Lyme arthritis by category of clinician suspicion (categorized into “unlikely” 1-3, “possible” 4-7, and “very likely” 8-10). We then compared the rate of joint procedures, parenteral antibiotics, and hospitalization by increasing clinician suspicion using Cochran-Armitage tests. We performed all analyses in R Studio (Version 4.5.2, Vienna, Austria).

## Results

We enrolled 1,582 children with monoarthritis evaluated for Lyme disease (median age 7 years, interquartile range 4-11 years). The majority of children were male (n=989; 63%) and presented during the peak Lyme season between June and October (n=829; 52%, **Supplemental Table 1**). The most affected joint was the knee (n=1,128; 71%) followed by the hip (n=249; 16%). Overall, 623 (39%) had Lyme arthritis, and 32 (2%) septic arthritis, with the majority attributed to *staphylococcus aureus* (n=15).

Of the 1,578 with clinician suspicion recorded (99.7% of study population), 607 (38%) were classified as unlikely, 663 (42%) possible and 308 (19%) very likely to have Lyme arthritis. Clinician suspicion had moderate discriminative ability for Lyme disease (AUC: 0.75, 95% CI: 0.72-0.77, **Figure 1**).^7^ Lyme arthritis was diagnosed in 20.4% (95% CI: 17.4%-23.8%) of children for whom clinicians estimated Lyme disease as “unlikely”, 40.1% (95% CI: 36.5%-43.9%) as “possible”, and 75.6% (95% CI: 70.6%-80.1%) as “very likely”. Children with higher clinician suspicion scores were less likely to receive parenteral antibiotics or to be hospitalized, although invasive procedure rates were similar (**Table 1**).

**Figure 1.**
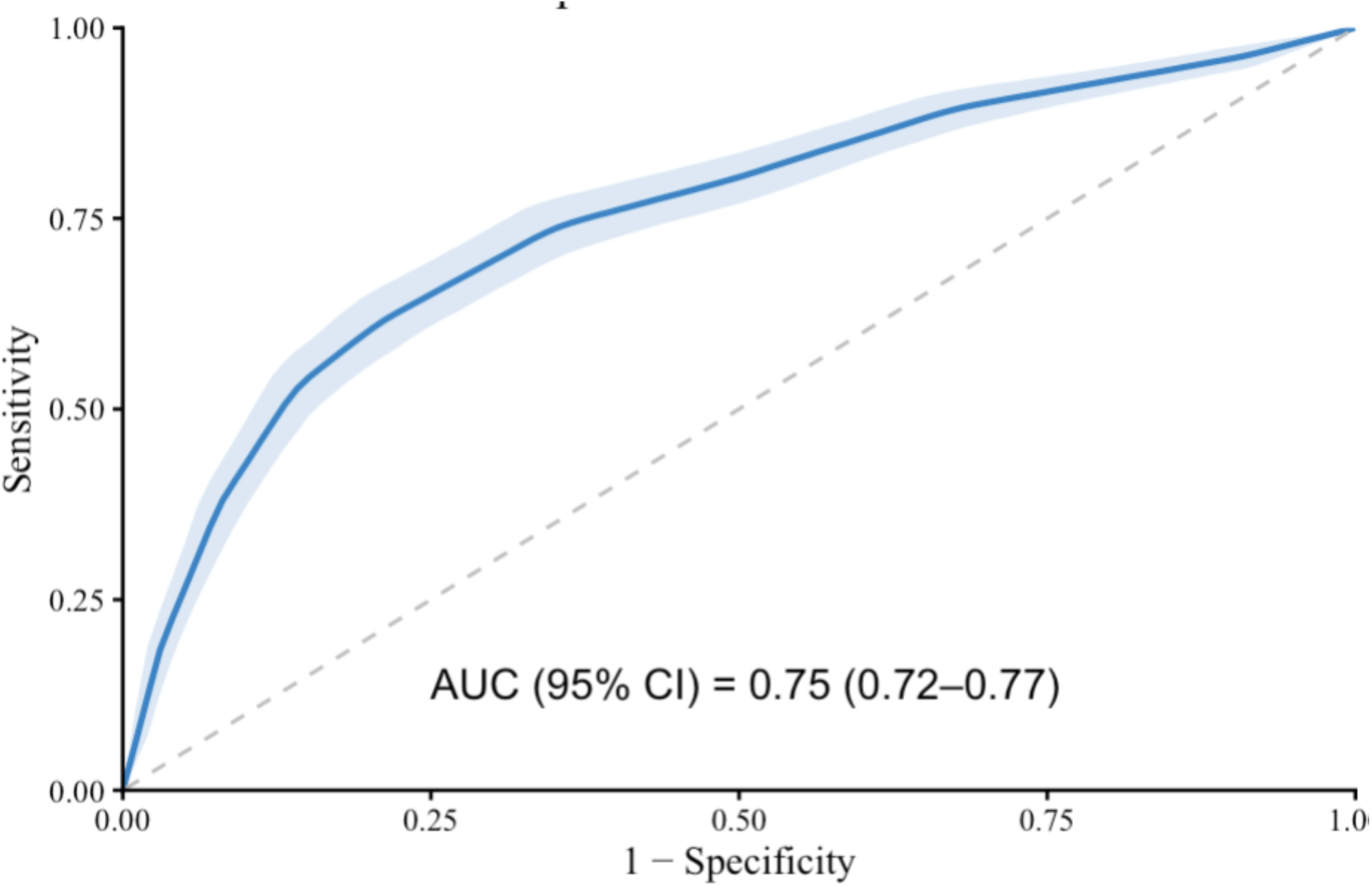
Receiver operating characteristics curve demonstrating the performance of clinician suspicion for Lyme disease to diagnose Lyme disease among children with monoarthritis. Area under the receiver operating characteristics curve (95% CI): 0.75 (0.72-0.77)

**Table 1:**
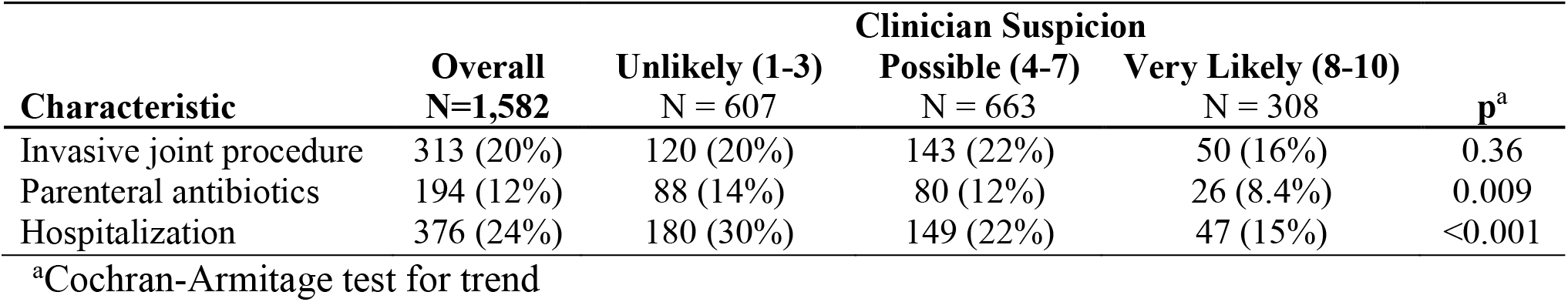
Clinical decision-making among children with monoarthritis, stratified by clinician suspicion for Lyme disease.

## Discussion

Our findings highlight the challenge of distinguishing Lyme from septic arthritis. Clinician suspicion had moderate discriminative ability and three-quarters of children categorized as “very likely” were ultimately diagnosed with Lyme arthritis. However, one in five children underwent an invasive joint procedure and many received parenteral antibiotics and/or hospitalization while awaiting results of Lyme disease testing. Although septic arthritis is rare and published tools identify children at low risk, the observed rates of invasive testing, irrespective of clinician suspicion, may reflect clinician discomfort in relying on clinician suspicion alone, given the risk of delayed diagnosis for septic arthritis.^2, 8, 9^

Our study has limitations. First, we enrolled children at centers in high incidence Lyme regions, so results may not be broadly generalizable to non-endemic regions. Second, we captured clinician suspicion at the time of initial evaluation before results of diagnostic testing were available. Management decisions regarding invasive procedures, treatment and ED disposition are typically made after review of diagnostic test which could have improved clinician accuracy.

In conclusion, more rapid diagnostic tools are needed to distinguish Lyme from septic arthritis. Validated clinical prediction models for Lyme arthritis could improve timely diagnosis and safely reduce invasive testing in children presenting with monoarthritis in Lyme-endemic regions.

## Data Availability

Due to the sensitivity of this clinical research, the dataset used to produce the aggregate data in this work is not publicly available.

## Abbreviations

ED: emergency department
ROC: receiver operating characteristics
AUC: area under the ROC curve
CI: confidence interval

