## Supplemental Table 1 for "Clinician Suspicion for Lyme Disease and Clinical Decision-Making in Children with Monoarthritis"

**Supplemental Table 1:** Cohort characteristics overall and stratified by clinician suspicion for Lyme disease

| Characteristic | Overall N = 1,582 | Clinician Suspicion (1-10) Unlikely (1-3) N = 607 | Possible (4-7) N = 663 | Very Likely (8-10) N = 308 | p[b] |
|---|---|---|---|---|---|
| ***Demographics*** | | | | | |
| Age, years | 7 (4, 11) | 8 (4, 12) | 7 (4, 10) | 7 (5, 10) | 0.088 |
| Male | 989 (63%) | 364 (60%) | 424 (64%) | 199 (65%) | 0.2 |
| Prior Lyme disease history[a] | 70 (4%) | 17 (3%) | 31 (5%) | 22 (7%) | 0.010 |
| ***Clinical Characteristics*** | | | | | |
| Lyme season (Jun-Oct) | 829 (52%) | 321 (53%) | 339 (51%) | 168 (55%) | 0.6 |
| Fever | 391 (25%) | 151 (25%) | 161 (24%) | 79 (26%) | 0.9 |
| Report of tick bite | 125 (8%) | 30 (5%) | 54 (8%) | 41 (13%) | <0.001 |
| Location | | | | | |
| Ankle | 116 (7%) | 68 (11%) | 38 (6%) | 8 (3%) | <0.001 |
| Elbow | 59 (4%) | 27 (4%) | 25 (4%) | 7 (2%) | 0.3 |
| Hip (pain or swelling) | 249 (16%) | 132 (22%) | 106 (16%) | 10 (3%) | <0.001 |
| Knee | 1,128 (71%) | 358 (59%) | 490 (74%) | 279 (91%) | <0.001 |
| Wrist | 20 (1%) | 15 (2.5%) | 4 (0.6%) | 1 (0.3%) | 0.004 |
| Other | 10 (0.6%) | 7 (1%) | 0 (0%) | 3 (1%) | 0.009 |
| ***Blood work***[c] | | | | | |
| White blood cell count | 9.4 (7.5, 11.4) | 9.3 (7.3, 11.6) | 9.5 (7.5, 11.4) | 9.4 (7.8, 11.2) | 0.7 |
| C-reactive protein | 1.01 (0.29, 3.00) | 0.71 (0.25, 2.80) | 1.02 (0.29, 2.87) | 1.81 (0.60, 3.74) | <0.001 |
| Erythrocyte sedimentation rate | 19 (10, 39) | 17 (8, 39) | 19 (10, 36) | 27 (14, 43) | <0.001 |

[a]Confirmed or possible

[b]Wilcoxon rank sum test for continuous variables, Chi-squared test or Fisher's exact (where cell counts <5) test for categorical variables

[c]Blood work missing for <15% of the study cohort
